# Outcomes from COVID-19 across the range of frailty: excess mortality in fitter older people

**DOI:** 10.1101/2020.05.22.20110486

**Authors:** Amy Miles, Thomas E Webb, Benjamin C Mcloughlin, Imran Mannan, Arshad Rather, Paul Knopp, Daniel Davis

## Abstract

**Purpose:** Our aim was to quantify the mortality from COVID-19 and identify any interactions with frailty and other demographic factors.

**Methods:** Hospitalised patients aged ≥70 were included, comparing COVID-19 cases with non-COVID-19 controls admitted over the same period. Frailty was prospectively measured and mortality ascertained through linkage with national and local statutory reports.

**Results:** In 217 COVID-19 cases and 160 controls, older age and South Asian ethnicity, though not socioeconomic position, were associated with higher mortality. For frailty, differences in effect size were evident between cases (HR 1.02, 95%CI 0.93-1.12) and controls (HR 1.99, 95%CI 1.46-2.72), with an interaction term (HR 0.51, 95%CI 0.37-0.71) in multivariable models.

**Conclusions:** Our findings suggest that (i) frailty is not a good discriminator of prognosis in COVID-19 and (ii) pathways to mortality may differ in fitter compared with frailer older patients.

**Key summary points:** *Aim:* To describe associations between frailty, ethnicity, socioeconomic position and mortality in a cohort of older patients presenting to hospital with COVID-19.

## Introduction

While it is clear that mortality from SARS-CoV-2 infection (COVID-19) increases with age [1–3], the association between frailty and mortality is not well understood [4, 5]. This relationship has clinical implications as the National Institute for Health and Care Excellence (NICE) guidelines in England and Wales recommend the integration of a frailty assessment into algorithms used to guide decisions including admission to critical care [6]. Furthermore, other demographic factors relevant to mortality such as ethnicity or socioeconomic position have yet to be comprehensively described in relation to COVID-19 [7, 8].

Our aim was to investigate the relationship between frailty, ethnicity, socioeconomic position and mortality in a cohort of older patients presenting to hospital with COVID-19, in order to: (i) quantify the mortality from COVID-19; (ii) identify any interactions with frailty and other demographic factors in this population.

## Methods

### Participants

Patients admitted to an urban teaching hospital aged ≥70 were included if they tested positive for SARS-CoV-2 by combined throat and high-nasal swab on reverse-transcriptase polymerase chain reaction or if there was high clinical suspicion (on the basis of clinical, imaging and laboratory results, as determined by specialist infectious diseases physicians) for COVID-19 up until 23^rd^ April 2020. During the pandemic, the index of suspicion for SARS-CoV-2 infection was very high, so each older person needing hospitalisation was systematically assessed for COVID-19. Therefore, the control group comprised patients aged ≥70 who had been admitted within the same time period who reliably did not have COVID-19.

### Outcome

Our primary outcome was all-cause mortality determined up until 13^th^ May 2020. Deaths occurring outside of hospital were captured through daily updates on the NHS Spine, a collection of local and national databases and systems containing demographic information.

### Exposures

Frailty was quantified by the Clinical Frailty Scale (CFS) score [9] capturing patients’ clinical state two weeks prior to admission. This was assessed prospectively by the admitting clinical team, though all scores were reviewed by specialist geriatricians. Socioeconomic position was estimated through the Index of Multiple Deprivation (IMD) (along with Health, Income, Education sub-indices) which is an ecological measure determined by home postcodes [10]. Ethnicity was self-reported in hospital administrative data.

### Statistical analysis

Differences in continuous or categorical variables were assessed using t-tests and χ^2^ tests respectively. Cox proportional hazards models estimated differences in survival between COVID-19 cases and non-COVID-19 controls. Frailty was considered a continuous variable, ethnicity was classified into *South Asian*, *Black*, *White*, *Mixed*, *Other*, and deciles of IMD (1 = most advantaged; 10 = most disadvantaged) and its sub-indices were used in univariable and multivariable models. Interactions between Clinical Frailty Score and COVID-19 status were assessed. Statistical significance was determined at p<0.05. Post-estimation procedures included Schoenfeld residuals to test heteroskedasticity. Stata 14.1 (StataCorp, Texas, USA) was used for all analyses.

## Results

A total of 217 COVID-19 cases and 160 non-COVID-19 controls were identified. In COVID-19 cases, the mean age was 80.0 (SD 6.8) (range 70 to 99) years (Table 1). The majority of cases were men (n=134, 62%) or of white ethnicity (n=138, 63%). There was a normal distribution of clinical frailty scores and the median Index of Multiple Deprivation decile was 4 (IQR 3, 6). There were no significant differences in age, ethnicity, Index of Multiple Deprivation decile, or Clinical Frailty Scale score between cases and controls (Table 1).

**Table 1.** Patient characteristics of study participants by COVID-19 status.

|  | **COVID-19 cases**<br>**(n = 217)** | **Controls**<br>**(n = 160)** | ***P*** |
| --- | --- | --- | --- |
| Age (SD) | 80.0 (7.6) | 81.4 (6.8) | 0.06 |
| Sex (%) M | 134 (61.8) | 78 (48.8) | 0.01 |
| Ethnicity (%) |  |  | 0.92 |
| South Asian | 16 (7.4) | 9 (5.6) |  |
| Black | 16 (7.4) | 15 (9.4) |  |
| White | 138 (63.6) | 103 (64.4) |  |
| Mixed | 19 (8.8) | 13 (8.1) |  |
| Unknown | 28 (12.9) | 20 (12.5) |  |
| Median IMD Decile (IQR) | 4 (3, 6) | 4 (2, 6) | 0.09 |
| Median Health and Disability Decile (IQR) | 6 (4, 8) | 6 (4, 8) | 0.14 |
| CFS (%) |  |  | 0.37 |
| 1 | 4 (1.9) | 2 (1.3) |  |
| 2 | 32 (14.9) | 14 (8.8) |  |
| 3 | 32 (14.9) | 24 (15.0) |  |
| 4 | 32 (14.9) | 36 (22.5) |  |
| 5 | 25 (11.6) | 25 (15.6) |  |
| 6 | 41 (19.1) | 26 (16.3) |  |
| 7 | 32 (14.9) | 24 (15.0) |  |
| 8 | 16 (7.4) | 9 (5.6) |  |
| 9 | 1 (0.5) | 0 (0.0) |  |
| Death | 111 (51.2) | 22 (13.8) | <0.01 |

Abbreviations: SD, standard deviation; IQR, interquartile range; CFS, Clinical Frailty Scale.

In univariable models, COVID-19, older age and South Asian ethnicity were associated with higher mortality, though no measure of socioeconomic position demonstrated any association (Table 2). For frailty, differences in effect size were evident between cases (HR 1.02, 95% CI 0.93 to 1.12, p=0.71) and controls (HR 1.99, 95% CI 1.46 to 2.72, p<0.01). In the multivariable model, these relationships remained consistent: age (HR 1.04, 95% CI 1.01 to 1.07, p<0.01), South Asian ethnicity (HR 1.13, 95% CI 1.13 to 3.51, p=0.02) (Table 2).

**Table 2.** Univariable and multivariable analysis of the effect of cohort characteristics on mortality in COVID-19

|  | Univariable models |  |  |  | Multivariable model |  |  |  |
| --- | --- | --- | --- | --- | --- | --- | --- | --- |
|  | HR | 95% CI |  | P | HR | 95% CI |  | P |
| Age | 1.03 | 1.01 | 1.06 | 0.01 | 1.04 | 1.01 | 1.07 | <0.01 |
| Sex | 1.41 | 0.99 | 2.01 | 0.06 | 1.39 | 0.96 | 2.01 | 0.08 |
| Ethnicity |  |  |  |  |  |  |  |  |
| South Asian | 2.08 | 1.20 | 3.6 | 0.01 | 2.00 | 1.13 | 3.51 | 0.02 |
| Black | 1.12 | 0.60 | 2.1 | 0.72 | 1.30 | 0.70 | 2.51 | 0.38 |
| White | [ref] |  |  |  | [ref] |  |  |  |
| Mixed | 1.00 | 0.53 | 1.89 | 0.99 | 0.93 | 0.49 | 1.77 | 0.83 |
| Unknown | 0.85 | 0.48 | 1.49 | 0.57 | 0.96 | 0.54 | 1.70 | 0.88 |
| IMD | 1.02 | 0.95 | 1.11 | 0.54 | 1.00 | 0.93 | 1.08 | 0.98 |
| Income | 1.04 | 0.97 | 1.11 | 0.27 |  |  |  |  |
| Education | 1.01 | 0.94 | 1.09 | 0.76 |  |  |  |  |
| Health | 1.02 | 0.95 | 1.09 | 0.56 |  |  |  |  |
| CFS | 1.12 | 1.02 | 1.23 | 0.02 | 1.88 | 1.37 | 2.59 | <0.01 |
| COVID-19 | 4.40 | 2.78 | 6.97 | <0.01 | 213 | 24.63 | 1842 | <0.01 |
| COVID-19 x<br>CFS interaction |  |  |  |  | 0.51 | 0.37 | 0.71 | <0.01 |

Abbreviations: HR, hazard ratio; CI, confidence interval; IMD, Index of Multiple Deprivation; CFS, Clinical Frailty Scale

The different associations with frailty according to COVID-19 status was confirmed by demonstrating an interaction term (HR 0.51, 95% CI 0.37 to 0.71, p<0.01). The coefficient direction suggests that mortality is *proportionally higher* in fitter patients. Estimated in this way, the overall mortality attributable to COVID-19 was extremely high in this population (HR 213, 95% CI 24.6 to 1841, p<0.01). When plotting mutually-adjusted survival curves, tertiles of CFS showed distinct trajectories in non-COVID-19 controls, not at all apparent in COVID-19 cases (Figure 1). In keeping with the interaction parameter, differences were most stark in fitter patients (CFS 1-3) but less so in frailer ones (CFS 7-9).

**Figure 1.**
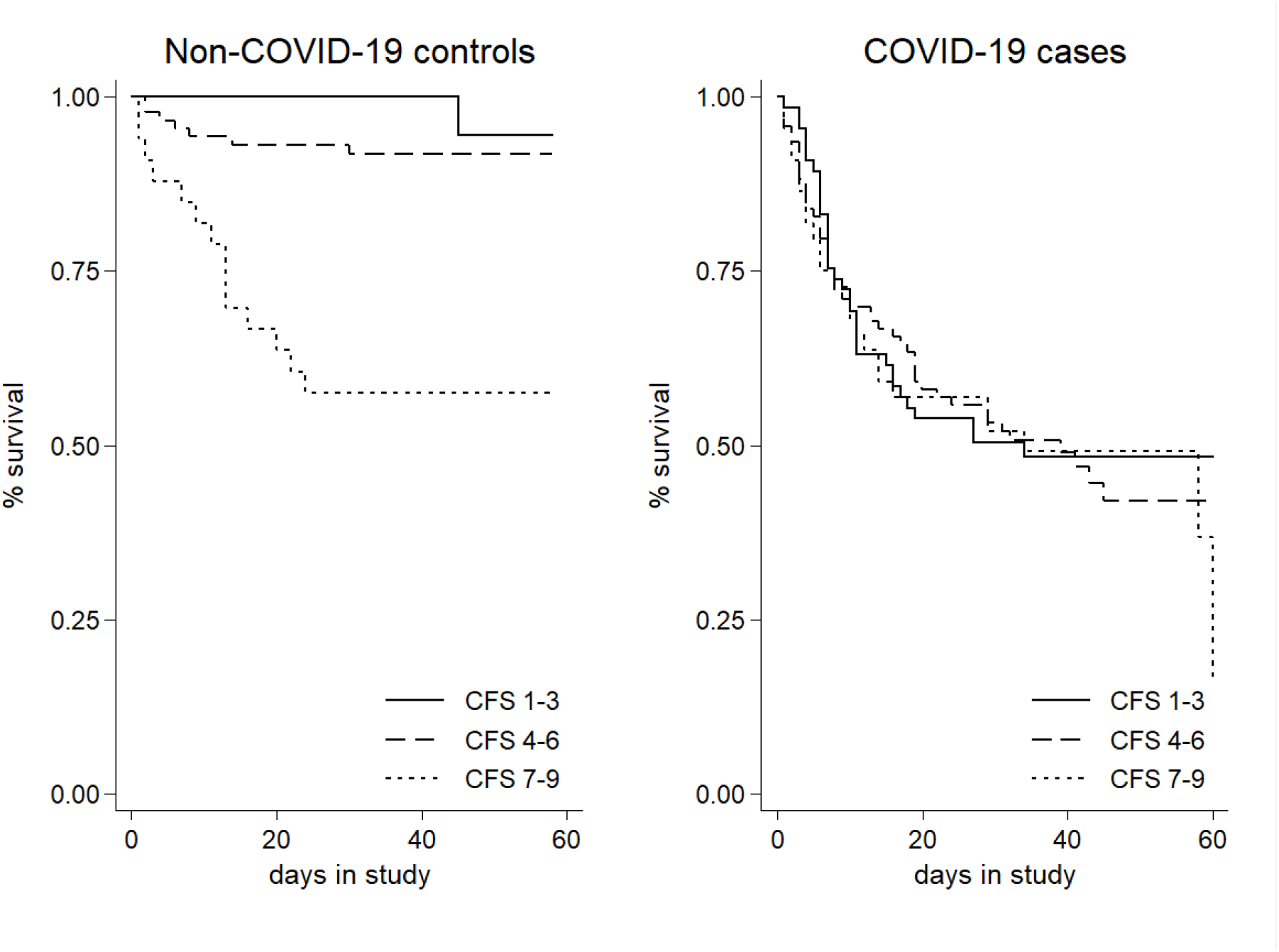
Kaplan-Meier curves showing 60 day survival by tertiles of Clinical Frailty Scale (CFS) and COVID-19 status

## Discussion

In this population of older admissions to a central London hospital, frailty did not appear to be associated with mortality rates after COVID-19. In addition, ecological measures of socioeconomic position were not associated with death, though there was some evidence of excess risk in South Asian compared with White populations. Associations with mortality in those with and without COVID-19 demonstrated much larger excess mortality in fitter, compared with frailer patients. Taken together, our findings suggest that (i) frailty is not a good discriminator of prognosis in COVID-19 and (ii) pathways to mortality may differ in fitter compared with frailer older patients.

Our results should be treated with caution. Data were collected from a single site, albeit a large teaching hospital at the first peak of the COVID-19 pandemic. For a proportion (21%), ethnicity was either mixed or undetermined, perhaps reflecting a casemix specific to London. Furthermore, ecological measures of socioeconomic position will be less reliable compared with individual factors, and the index of multiple deprivation may not relate to health outcomes as well in London residents [11]. Our results are only applicable to hospitalised patients, and some selection bias might arise from different indications for self-presenting to secondary care in COVID-19 patients versus those without respiratory symptoms (our controls). Nonetheless, our data have the advantage of specialist assessments of COVID-19 status and frailty, as well as accurate statutory reporting of dates of death.

These findings add to emerging reports quantifying the relationship between frailty and mortality in COVID-19. In another London hospitalised cohort, crude deaths in COVID-19 were higher in patients who were frailer (median Clinical Frailty Scale score of 5 versus 4, p=0.01) [12]. Other UK case series have shown that patients who died without ventilatory support had a median Clinical Frailty Scale score of 7 [13]. To date, most studies are describing mortality without reference to a contemporaneous non-COVID-19 population, which would obscure the interaction apparent in our data. In this respect, our findings are most consistent with comparable data from Leicester which also show no association between frailty and mortality in COVID-19 (Simon Conroy, personal correspondence).

Our findings have two major implications. Firstly, if frailty states in COVID-19 are not associated with mortality, then this has only limited value as a consideration in older people who may require ventilatory support. This is in contrast to the central NICE guidance that recommends a frailty assessment as the first step in the assessment for critical care. Secondly, an interaction between COVID-19 and frailty implies that different pathways to death could be at play. In general, the pathophysiology described in COVID-19 patients in critical care indicates substantial immune hyperactivation [14]. However, given survival in the CFS range 7-9 was similar in cases and controls, this may reflect death from COVID-19 is occurring in the same way as for other common illness and immune hyperactivation is unlikely to be a significant feature in this group. One might speculate that older people with frailty have pre-existing immunesenescence such that they are unable to mount excess immune responses and may be otherwise by dying from the direct effects of viral infection.

While COVID-19 clearly confers substantial mortality in older people, we show that this risk may arise for different reasons depending on pre-morbid frailty. Further work should consider other outcomes after COVID-19, particularly cognitive and physical function. If baseline frailty and associated immunesenscence influences the subsequent inflammatory response, this hints that different therapeutic strategies might be needed across the spectrum of frailty.

## Data Availability

Data available on request

## Findings

Frailty did not appear to be associated with mortality rates after COVID-19, though an interaction was evident indicating much larger excess mortality in fitter, compared with frailer patients.

## Message

Frailty may not be a good measure of prognosis in COVID-19 and different mechanisms may underlie pathways to death depending on pre-morbid frailty.

## Declarations

### Funding

Daniel Davis is funded through a Wellcome Intermediate Clinical Fellowship (WT107467).

### Conflicts of interest

The authors declare that they have no conflict of interest.

### Ethics approval

These analyses were conducted as part of a service evaluation project and individual consent was not necessary as determined by the NHS Health Research Authority.

### Availability of data and material

on request

### Code availability

on request

### Authors' contributions

AM, TW, BM, PK collected the primary data. DD undertook the statistical analyses and had oversight of the project. AM and TW drafted the first version of the manuscript. All authors contributed to revision and intellectual content of the final submission

## Notes

### Competing Interest Statement

The authors have declared no competing interest.

